# ACTH-independent hypercortisolism across the cardiometabolic spectrum

**DOI:** 10.64898/2026.09.11.26362842

**Authors:** Andrew J. Newman, Sanan Mahrokhian, Isabelle Hanna, Cheng-Hsuan Tsai, Stéfanie Parisien-La Salle, Manaporn Payanundana, Richard J. Auchus, Jenifer M. Brown, Anand Vaidya

## Abstract

**Background:** Accumulating evidence suggests that the prevalence of hypercortisolism in patients with cardiometabolic risk factors is much higher than previously thought. This study aimed to evaluate the prevalence of ACTH-independent hypercortisolism across the spectrum of cardiometabolic risk.

**Methods:** Participants were prospectively recruited into three cohorts to undergo protocolized assessment of adrenal physiology:1) normotensive participants; 2) participants with hypertension and obesity, but without diabetes; 3) participants with diabetes and overweight/obesity. All participants (n=216) underwent overnight 1 mg dexamethasone suppression testing followed by cosyntropin stimulation test, and 24-hour urine free cortisol (UFC) testing. ACTH-independent hypercortisolism was defined as post-dexamethasone serum cortisol >1.8 μg/dL (50 nmol/L) with a concomitant post-dexamethasone ACTH ≤10 pg/mL (2.2 pmol/L).

**Result:** 16 of 216 (7.4%) participants were found to have ACTH-independent hypercortisolism, including 5.6% (4/71) in the normotensive cohort, 6.2% (5/81) in the hypertension-obesity cohort and 10.9% (7/64) in the diabetes-obesity cohort. Age, body mass index, hemoglobin A1c, blood pressure, renal function, morning ACTH, and 24-hour UFC were similar among those with and without ACTH-independent hypercortisolism. Following cosyntropin stimulation, those with ACTH-independent hypercortisolism had higher stimulated cortisol levels (greater ACTH-dependent responses) when compared to those without ACTH-independent hypercortisolism: 25.8±4.5 versus 20.6±4.1 μg/dL (711±124 v. 568±113 nmol/L) (*P*<0.001).

**Conclusions:** In this prospective study, the prevalence of ACTH-independent hypercortisolism paralleled the burden of cardiometabolic risk features. ACTH-independent hypercortisolism was associated with greater ACTH-stimulable cortisol production, identifying a unique biochemical phenotype. These findings reveal new insights into cortisol pathophysiology as a function of ACTH and cardiometabolic risk profiles.

## INTRODUCTION

Hypercortisolism in its severest form, Cushing syndrome, is well-established as driver of cardiometabolic risk and adverse outcomes such as obesity, diabetes mellitus, hypertension, dyslipidemia, and cardiovascular disease.^1,2^ Accumulating evidence supports associations between milder degrees of hypercortisolism, especially as mild autonomous cortisol secretion (MACS), a form of adrenocorticotropic hormone (ACTH)-independent adrenal hypercortisolism, and the same cardiometabolic risk factors.^3,4^

Recently, large cohort studies have demonstrated remarkably high rates of hypercortisolism, defined as dexamethasone non-suppressible cortisol > 1.8 μg/dL after 1 mg overnight dexamethasone suppression test (DST), in populations with difficult-to-treat diabetes (CATALYST)^5^ and resistant hypertension (MOMENTUM).^6^ In both studies, approximately one quarter of participants had a positive DST, implicating a possible role for hypercortisolism in the pathogenesis of difficult to control diabetes and hypertension. Notably, treatment of this hypercortisolism with glucocorticoid-receptor antagonism resulted in improvements in glycemic control in the CATALYST trial.

Whether this surprisingly high prevalence of hypercortisolism was driven by ACTH, or independent of ACTH, remains incompletely characterized. Further, the prevalence of hypercortisolism in populations with less severe manifestations of cardiometabolic disease has not been thoroughly established. In this study, we aimed to evaluate the frequency of ACTH-independent hypercortisolism using protocolized testing in three cohorts with low, medium, and high cardiometabolic risk.

## METHODS

### Study cohorts

Participants were prospectively recruited from the greater Boston, MA, USA, area into three cohorts to undergo phenotyping procedures to evaluate adrenal physiology.

The low risk cohort was a normotension (NTN) cohort,^7^ including n=74 participants who did not carry a diagnosis of hypertension but were at elevated risk of developing hypertension. They were age 35-70 years, with blood pressure of 115 – 135 / 70 – 85 mmHg with one or two of the following risk factors (depending on baseline blood pressure): body mass index (BMI) ≥ 25 kg/m^2^; first-degree family history of hypertension diagnosed before age 60; diabetes mellitus with hemoglobin A1c ≤ 9%.

The medium risk cohort was a hypertension-obesity (HTN-OB) cohort,^8^ including n=84 participants who had overweight or obesity with additional cardiometabolic risk factors, but excluding overt diabetes. They were age 18 – 70 years, with BMI ≥ 30 kg/m^2^ and at least one of the following additional risk factors or BMI 25 – 29.9 kg/m^2^ and at least two additional risk factors: established hypertension with screening BP ≤ 160 / 100 mmHg on treatment or not; impaired fasting glucose (100-125 mg/dL) or pre-diabetes by A1c (5.7 – 6.4%) but not diabetes; or dyslipidemia (fasting triglycerides > 150 mg/dL and HDL < 40 mg/dL [men] or HDL < 50 mg/dL [women]).

The high risk cohort was a diabetes-obesity cohort (DM-OB), including n=72 participants who had type 2 diabetes (T2DM) with screening A1c < 9%. Participants had either established CKD3a with estimated glomerular filtration rate (eGFR) 45-59 mL/min/1.73 m^2^, or were at-risk for CKD with eGFR 60-89 mL/min/1.73 m^2^ plus at least one of the following: moderate albuminuria (30-300 mg/g creatinine); diagnosis of hypertension and/or treatment with anti- hypertensive medications; or BMI ≥ 30 kg/m^2^.

In all three cohorts, individuals with known adrenal disease (including adrenal insufficiency and Cushing syndrome) and oral glucocorticoid use were excluded. In the two cohorts that allowed treated hypertension (HTN-OB and DM-OB), individuals using mineralocorticoid receptor antagonists were excluded, and those using ACE inhibitors or angiotensin receptor blockers were washed off these medications for 3-7 days prior to study procedures, with substitution with amlodipine or doxazosin if needed.

### Study procedures

Participants underwent a standard, low-dose dexamethasone suppression test (DST). They took dexamethasone 1 mg by mouth at 11 pm followed by fasting, upright, seated venipuncture at 8-9 AM the next morning. A post-DST cortisol > 1.8 μg/dL (50 nmol/L) with post-DST ACTH ≤ 10 pg/mL (2.2 pmol/L) was interpreted as functional validation consistent with ACTH-independent hypercortisolism. Participants were excluded from the analysis if post-DST ACTH > 10 pg/mL (2.2 pmol/L) to ensure evaluation of only ACTH-independent cortisol production. The DST was immediately followed by a standard cosyntropin stimulation test with cosyntropin 250 μg intravenous bolus followed by seated venipuncture 60 minutes later.

Serum cortisol, ACTH, plasma renin activity, and aldosterone were also measured on a separate day at 8-9 AM, while seated, following an oral sodium loading test (OSLT). Plasma renin and aldosterone values were included only if the 24 hour urine sodium excretion exceeded 3.5 g/24h (150 mmol/24h). These results were used to establish 8 AM cortisol and ACTH values and to evaluate the renin-angiotensin-aldosterone system.

### Laboratory Assays

ACTH was measured by ELISA (ALPCO; Cat# 21-ACTHU-E01, RRID:AB_3714847). Serum cortisol was measured by immunoassay (Beckman-Coulter; Cat# 33600, RRID:AB_2802133). Urine free cortisol was measured by liquid chromatography-tandem mass spectrometry at the Mass General Brigham Research Assay Core. Plasma aldosterone was measured by immunoassay (IBL-America; Cat# IB79134, RRID:AB_2813725). Plasma renin activity was measured by immunoassay of angiotensin I in plasma after incubation for 3 h at 37 °C (IBL-America Cat# IB59131, RRID: AB_3532145).

### Statistical methods

Data are reported as mean ± standard deviation for normally distributed variables, median (interquartile range) for non-normally distributed variables, and frequency (percent) for categorical variables. Hypothesis testing of normally distributed variables was performed with Student’s t-test; of non-normally distributed variables with the Wilcoxon rank-sum test; and of categorical variables with the Fisher’s Exact test. Analyses were performed with R version 4.5.2 in RStudio version 2026.07.1+147.

### Ethics & Oversight

All participants provided written informed consent and study protocols were approved and monitored by the Mass General Brigham Institutional Review Board (Protocols #2018P000257, 2020P002311, and 2023P003466) in compliance with the Declaration of Helsinki.

## RESULTS

### Characteristics of the study cohorts

Demographic characteristics of study participants are listed in **Table 1**. 216 participants had post-DST ACTH ≤ 10 pg/mL [2.2 pmol/L] and thus were included in the analysis, with 71 in the NTN cohort, 81 in the HTN-OB cohort, and 64 in the DM-OB cohort. There were 14 participants across the three cohorts that were excluded because their post-DST ACTH was > 10 pg/mL (2.2 pmol/L) indicating non-suppressibility of ACTH and/or unexpected issues with dexamethasone administration or metabolism.

**Table 1.** Characteristics of all participants together and the three cohorts separately. BMI: body mass index; SBP: systolic blood pressure; DBP: diastolic blood pressure; eGFR: estimated glomerular filtration rate; A1c: Hemoglobin A1c; ACTH: adrenocorticotropic hormone.

|  | All | Cohort |  |  |
| --- | --- | --- | --- | --- |
|  |  | NTN | HTN-OB | DM-OB |
| N | 216 | 71 | 81 | 64 |
| Age (y) | 56.3 ± 11.6 | 49.3 ± 10.3 | 56.2 ± 9.6 | 66.9 ± 8.1 |
| # Female (%) | 126 (58.3) | 38 (53.5) | 56 (69.1) | 32 (50) |
| # White (%) | 180 (83.3) | 59 (83.1) | 68 (84) | 53 (82.8) |
| BMI (kg/m<sup>2</sup>) | 32.3 ± 5.7 | 29.5 ± 4.8 | 35.2 ± 5.4 | 31.7 ± 5.4 |
| SBP (mmHg) | 124.8 ± 11.3 | 121.5 ± 8 | 129.8 ± 11.6 | 122.0 ± 12.0 |
| DBP (mmHg) | 77.6 ± 8.9 | 77.2 ± 5.3 | 82.4 ± 8.5 | 71.9 ± 9.2 |
| eGFR<br>(mL/min/1.73 m<sup>2</sup>) | 83.5 ± 15.2 | 90.3 ± 12.1 | 87 ± 13.9 | 71.4 ± 12.7 |
| A1c (%) | 5.7 ± 0.7 | 5.4 ± 0.4 | 5.4 ± 0.4 | 6.5 ± 0.7 |
| Baseline AM cortisol<br>(µg/dL) | 8.8(6.7-11.4) | 8.5(7.1-11.4) | 8.2(6.3-10.4) | 9.5(7.3-12.2) |
| Baseline AM ACTH<br>(pg/mL) | 18.1(13.0-26.5) | 17.6(10.3-31.3) | 18(13.6-22.7) | 20.8(14.1-29.3) |
| 24h Urine Free<br>Cortisol (µg /24 h) | 21.4(13.9-33.5) | 22.2(13.2-32.9) | 21.4(15.2-33.1) | 21.3(12.0-37.5) |

Mean age was 56.3 ± 11.6 y overall, increasing from 49.3 ± 10.3 y in the NTN cohort to 66.9 ± 8.1 y in the DM-OB cohort. Participants were evenly split between male and female in the NTN and DM-OB cohorts, but slightly more females enrolled in the HTN-OB cohort. 83-84% of participants self-identified as white across all cohorts, similar to the Greater Boston, MA, area. By design, the mean BMI and on-treatment systolic and diastolic blood pressures were greatest in the HTN-OB cohort, while eGFR was lowest and on-treatment hemoglobin A1c highest in the DM-OB cohort. Baseline morning cortisol, ACTH, and 24-hour urine free cortisol were similar across cohorts.

### Frequency of ACTH-independent hypercortisolism across cohorts

16 of 216 (7.4%) participants across all three cohorts had post-DST cortisol > 1.8 μg/dL (50 nmol/L). In the NTN cohort, 4 of 71 (5.6%) participants had post-DST cortisol > 1.8 μg/dL (50 nmol/L), while 5 of 81 (6.2%) in the HTN-OB cohort and 7 of 64 (10.9%) in the DM-OB cohort had post-DST cortisol > 1.8 μg/dL (50 nmol/L) (**Fig. 1A**). **Fig. 1B** shows all post-DST cortisol values across all cohorts in rank order, colored by cohort, to visually demonstrate the higher post-DST cortisol levels in parallel with the transition in cohort risk factors.

**Figure 1.**
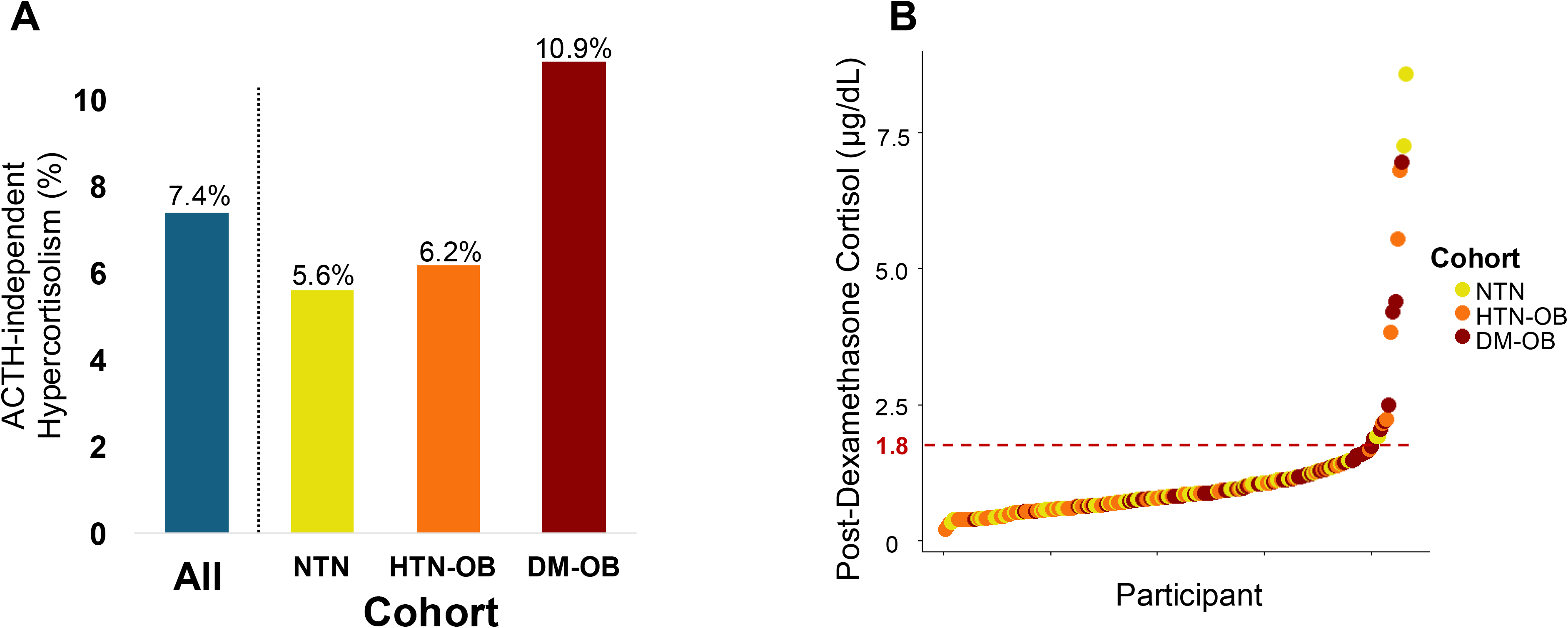
Frequency of ACTH-independent hypercortisolism across the three cohorts. **A.** Percentage of participants with ACTH-independent hypercortisolism across all cohorts combined and the three cohorts separately. **B.** All post-dexamethasone cortisol values colored by cohort. 1.8 μg/dL is highlighted as the conventional threshold for abnormal post-dexamethasone cortisol.

### Characteristics of those with and without ACTH-independent hypercortisolism

The 16 participants with ACTH-independent hypercortisolism, compared with the 200 participants without ACTH-independent hypercortisolism, had similar age, BMI, on-treatment systolic and diastolic blood pressure and eGFR (**Table 2**). Though there were proportionally fewer female and white participants among those with than without ACTH-independent hypercortisolism, these differences were not significant (P = 0.29 and 0.31, respectively). 8 AM morning cortisol (**Fig. 2A**) and 24h urine free cortisol (**Fig. 2C**) were numerically but not statistically significantly higher among those with ACTH-independent hypercortisolism, while 8 AM morning ACTH (**Fig. 2B**) did not differ. The mean 60 min post-cosyntropin cortisol among those with ACTH-independent hypercortisolism was 25.8 ± 4.5 μg/dL (711 ± 124 nmol/L), and significantly higher compared with 20.6 ± 4.1 μg/dL (568±113 nmol/L) among those without ACTH-independent hypercortisolism (P < 0.001; **Fig. 3A**). In **Fig. 3B**, all post-cosyntropin cortisol results are displayed, with those with ACTH-independent hypercortisolism clustering toward the higher values of ACTH-dependent cortisol production. Within each cohort, those with ACTH-independent hypercortisolism had 60 min post-cosyntropin cortisol above the cohort median except for just 2 of the 6 in the DM-OB group who completed the cosyntropin stimulation (**Fig. 3C**).

**Figure 2.**
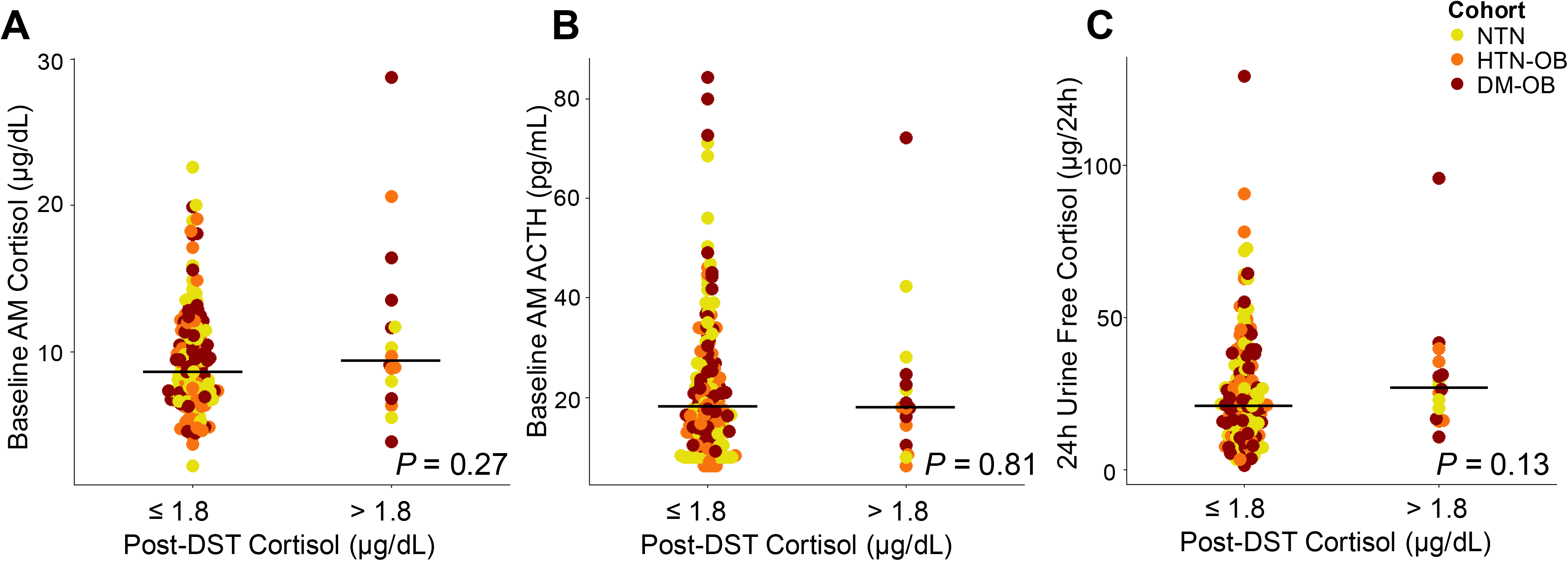
Comparison of **A.** 8 AM cortisol; **B.** 8 AM ACTH; and **C.** 24-hour urine free cortisol between those with and without ACTH-independent hypercortisolism, colored by cohort. Bars represent the median values.

**Figure 3.**
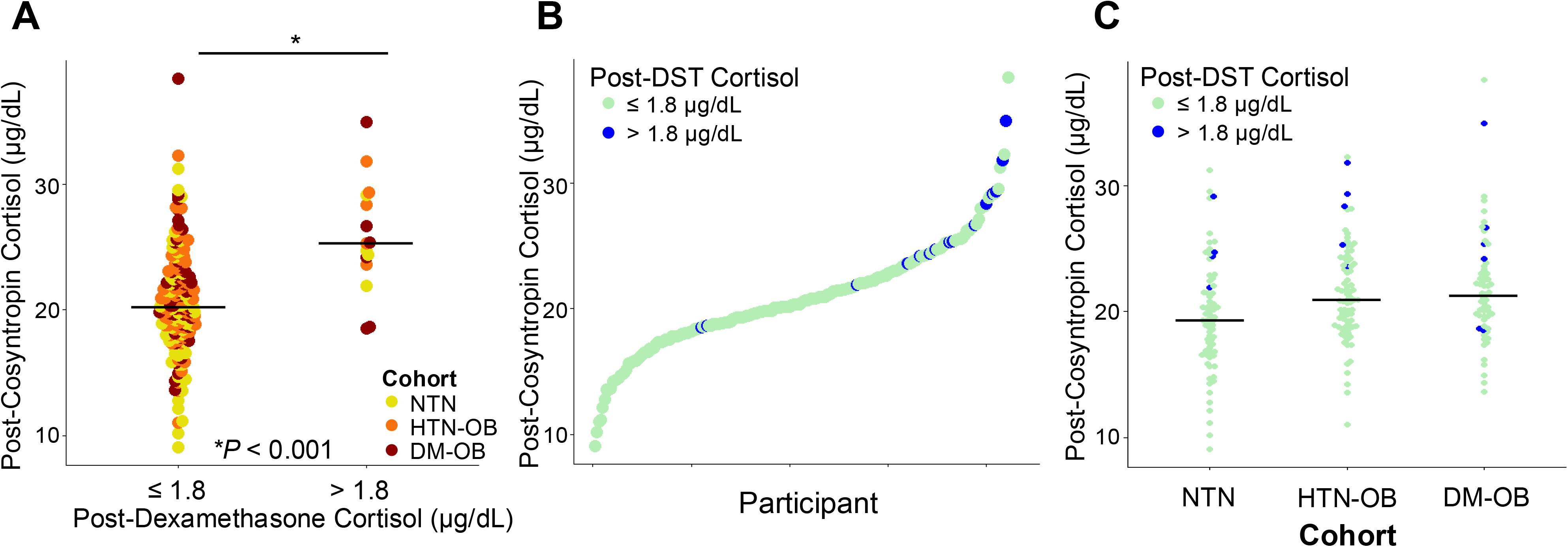
Post-cosyntropin cortisol values shown **A.** separated by post-dexamethasone cortisol (i.e., the presence or absence of ACTH-independent hypercortisolism) colored by cohort; **B.** across all participants, colored according to post-dexamethasone cortisol level; and **C.** by cohort, colored according to post-dexamethasone cortisol level.

**Table 2.** Characteristics of participants with post-dexamethasone cortisol ≤ 1.8 μg/dL (*without* ACTH-independent hypercortisolism) compared with those with post-dexamethasone cortisol > 1.8 μg/dL (*with* ACTH-independent hypercortisolism). BMI: body mass index; SBP: systolic blood pressure; DBP: diastolic blood pressure; eGFR: estimated glomerular filtration rate; A1c: Hemoglobin A1c; ACTH: adrenocorticotropic hormone. CST: cosyntropin stimulation test; PRA: plasma renin activity; ARR: aldosterone-to-renin ratio. *PRA, aldosterone and ARR analyses include only participants who had 24-hour urine sodium > 150 mmol/24h (N = 167 for post-DST cortisol ≤ 1.8 μg/dL group; N = 14 for post-DST cortisol > 1.8 μg/dL group).

| | post-DST Cortisol ($\mu\text{g/dL}$) | | P-value |
| --- | --- | --- | --- |
| | $\leq 1.8$ | $> 1.8$ | |
| N | 200 | 16 |  |
| Age (y) | 56.9 ± 11.8 | 58.8 ± 11.2 | 0.54 |
| # Female (%) | 119(59.5) | 7(43.8) | 0.29 |
| # White (%) | 168(84) | 12(75) | 0.31 |
| BMI ($\text{kg/m}^2$) | 32.3 ± 5.7 | 32.1 ± 5.9 | 0.93 |
| SBP (mmHg) | 124.6 ± 11.2 | 126.4 ± 13.4 | 0.62 |
| DBP (mmHg) | 77.8 ± 9.0 | 74.2 ± 7.7 | 0.09 |
| eGFR ($\text{mL/min/1.73 m}^2$) | 83.5 ± 15.2 | 83.3 ± 16.1 | 0.97 |
| A1c (%) | 5.7 ± 0.7 | 6.1 ± 1.0 | 0.10 |
| Baseline AM cortisol ($\mu\text{g/dL}$) | 8.7(6.7-11.2) | 9.4(7.4-12.7) | 0.27 |
| Baseline AM ACTH ($\text{pg/mL}$) | 18.3(13.0-26.6) | 18.0(12.5-23.6) | 0.81 |
| Post-CST cortisol ($\mu\text{g/dL}$) | 20.6 ± 4.1 | 25.8 ± 4.5 | $< 0.001$ |
| 24h Urine Free Cortisol ($\mu\text{g/24 h}$) | 21.2(12.9-33.5) | 27.0(18.5-33.4) | 0.13 |
| PRA ($\text{ng/mL/h}$) | 0.6(0.3-1.2) | 0.25(0.1-0.7) | $< 0.05$ |
| Aldosterone ($\text{ng/dL}$) | 14.0(8.7-20.7) | 11.2(8.4-19.3) | 0.68 |
| ARR ($[\text{ng/dL}]/[\text{ng/mL/h}]$) | 25.6(13.9-43.6) | 57.4(16.1-113.7) | $< 0.05$ |

Participants with ACTH-independent hypercortisolism had lower plasma renin activity than those without ACTH-independent hypercortisolism (0.25 (0.1-0.7) ng/mL/h v. 0.6 (0.3-1.2) ng/mL/h; P = 0.031), but similar plasma aldosterone concentrations (11.2 (8.4-19.3) ng/dL v. 14.0 (8.7-20.7) ng/dL; P = 0.68), and thus higher aldosterone-to-renin ratio (57.4 (16.1-113.7) ng/dL per ng/mL/h v. 25.6 (13.9-43.6) ng/dL per ng/mL/h; P = 0.035) following an OSLT.

## DISCUSSION

We found increasing frequency of ACTH-independent hypercortisolism across three prospectively recruited cohorts of asymptomatic research volunteers reflecting the cardiometabolic risk spectrum. Rates of ACTH-independent hypercortisolism ranged from 5.6% in the lowest risk NTN cohort to 10.9% in the highest risk DM-OB cohort. Those with and without ACTH-independent hypercortisolism did not differ in non-suppressed morning cortisol or ACTH, nor in total daily cortisol production as determined by urine free cortisol. Intriguingly, those with ACTH-independent hypercortisolism had higher cortisol production in response to exogenous ACTH administration, suggesting a state of adrenal hyperstimulability in response to ACTH. Further, these participants with ACTH-independent hypercortisolism also had lower renin with similar aldosterone production, potentially indicating additional renin suppression by excess cortisol exerting mineralocorticoid effects.

Several previous studies have examined the rate of hypercortisolism in patients with conditions exacerbated by cortisol excess. These studies have generally examined dexamethasone non-suppressible cortisol, defined as post-dexamethasone cortisol > 1.8 μg/dL without respect to ACTH, rather than ACTH-independent hypercortisolism specifically. The frequency of dexamethasone non-suppressible cortisol in patients with T2DM has been studied since at least the 1980s,^9^ when Hudson, et al., found that a higher rate of non-suppressed afternoon cortisol after nighttime dexamethasone among people with T2DM compared with those without T2DM. Catargi, et al., reported positive DST results in 26% of participants with T2DM and A1c > 8% and without specific signs or symptoms of hypercortisolism, with 5.5% having Cushing syndrome (defined as elevated cortisol on two types of tests plus a candidate tumor on imaging). The 15% with only an abnormal DST were categorized as false-positives and “a milder degree of occult [Cushing syndrome] is unlikely.”^10^ Similarly, Newsome, et al., found that 31 of 171 (18%) patients with T2DM and BMI > 25 kg/m^2^ had post-DST cortisol > 1.8 μg/dL (50 nmol/L).^11^ However, because just one patient also had a true elevated urine free cortisol, the other positive DST results were interpreted as false-positives. This interpretation in both studies reflects the underlying paradigm that the DST is a screening test for overt Cushing syndrome, the diagnosis of which requires elevated results on at least one other type of cortisol test.^12^ Newsome and colleagues expressed concern about the high rate of “false-positive DST results … and that the results were a continuum throughout the patients with diabetes, without any clear-cut point to distinguish normal from abnormal.”^11^ In our study, we identified a similar continuum, though the highest risk cohort seemed to cluster toward the higher end of the “normal” range.

Because these prior studies did not measure a marker of test adequacy, such as dexamethasone or post-dexamethasone ACTH level, results must be interpreted with caution. Admittedly post-dexamethasone ACTH level is useful as a marker of test adequacy only when considering ACTH-independent hypercortisolism alone, not all-cause Cushing syndrome. We relied on ACTH suppression post-dexamethasone as a physiologic and functional confirmation of dexamethasone effect to provide confidence that cortisol production was truly independent of ACTH.

Multiple studies have evaluated the frequency of dexamethasone non-suppressible hypercortisolism in obesity. Prevalence estimates vary, from 1%^13,14^ −9%,^15,16^ with some finding a positive correlation between BMI and frequency of hypercortisolism,^13^ and some finding no relationship.^17^ These studies suffer from similar limitations as the diabetes studies above, especially in labeling as “false positives” those cases with non-suppressed cortisol in the absence of another type of test corroborating hypercortisolism.

Fewer prior studies have evaluated hypercortisolism in patients with resistant hypertension. In one study, of 423 adults with resistant hypertension, 26.5% had post-DST cortisol > 1.8 μg/dL (50 nmol/L), though dexamethasone level and post-DST ACTH were not measured.^18^ Overall, 8% had hypercortisolism confirmed by a second cortisol test.

The CATALYST-1^5^ and MOMENTUM^6^ studies were large (N > 1000), multicenter studies enrolling participants with difficult-to-control diabetes (CATALYST-1) or resistant hypertension (MOMENTUM), who underwent 1 mg DST with measurement of dexamethasone level, Patients with abnormal post-DST cortisol > 1.8 μg/dL (50 nmol/L) had measurement of non-suppressed morning ACTH, DHEAS and cortisol to assess ACTH dependence, plus adrenal imaging. In CATALYST-1, 23.8% of participants had a positive DST (defined as post-dexamethasone cortisol > 1.8 μg/dL with dexamethasone level ≥ 140 ng/dL irrespective of ACTH), while in MOMENTUM, 27.3% of participants had a positive DST. Compared to our study cohorts, the frequency of positive DST in CATALYST-1 and MOMENTUM were 2-4 fold higher, corresponding to a substantially greater cardiometabolic disease severity. This supports the notion that severity of cardiometabolic risk burden may be associated with higher risk for hypercortisolism.

In CATALYST-1, the mean 8 AM ACTH and DHEA-S values were normal.^5^ This is in agreement with our finding that those with and without ACTH-independent hypercortisolism did not differ in 8 AM ACTH and cortisol. It is likely that the mild excess cortisol is not adequate to exert negative feedback on the hypothalamus and pituitary.

Notably, in CATALYST-1, just 34.7% of participants with hypercortisolism who underwent imaging had an adrenal abnormality that could explain hypercortisolism from an ACTH-independent source, while the remaining 65% are unexplained. Future studies should include post-dexamethasone ACTH to exclude overactivation of the hypothalamic-pituitary unit by the physiologic stress of cardiometabolic risk factors, as in non-neoplastic hypercortisolism.^19^ This would evaluate the possibility of reverse causation, i.e. that dexamethasone non-suppressible hypercortisolism is due to, rather than a cause of, cardiometabolic risk factors.

The relationship between post-DST cortisol and ACTH stimulability was described in small studies in the 1980s examining patients with depression (many of whom do not suppress cortisol with the standard 1 mg DST). Results varied, with some studies demonstrating a higher ACTH stimulated cortisol in patients who did not suppress cortisol with dexamethasone compared with those who did suppress;^20,21^ one study found that this difference disappeared after treatment of depression.^22^ Participants were not evaluated for active mood disorders in our study, and the generalizability of these results are uncertain. However, the hyper-stimulability by ACTH may indicate that ACTH-independent hypercortisolism may also portend greater stress-induced cortisol exposures, thereby providing an additional mechanism by which ACTH-independent hypercortisolism contributes to cardiometabolic disease, or vice-versa. Analogously to ACTH-independent hypercortisolism, we have shown that in renin-independent hyperaldosteronism (i.e. primary aldosteronism), people with less suppressible aldosterone have greater aldosterone stimulability by ACTH.^7,23^

### Strengths and limitations

Strengths of our study include prospective recruitment of participants with more common cardiometabolic risk profiles, who were not selected for risk of hypercortisolism, as well as thorough physiologic testing, including comprehensive manipulation of the hypothalamic-pituitary-adrenal axis and renin-angiotensin-aldosterone system. However, limitations should be recognized. Due to the small sample sizes, subtler differences between cohorts could not be determined. We did not conduct imaging to identify the source of excess cortisol. While we did not measure dexamethasone levels, our study considered a more physiologic read-out of ACTH-independent hypercortisolism, appropriately suppressed ACTH following the administration of dexamethasone.

Notably, participants’ age increased across the cohorts in parallel to the cohorts’ cardiometabolic risk burden. Age is an important potential confounder for which our study was underpowered to adjust, though the prevalence of most cardiometabolic risk factors and the prevalence of adrenal nodules that are the source of ACTH-independent hypercortisolism in some patients increase with age.^24,25^ It may be impossible to disentangle the independent influence of age as a mediator or bystander of the relationship between hypercortisolism and cardiometabolic burden since it is essentially collinear with both the exposure and the outcome in every population.

### Future directions

Interest is growing in mild hypercortisolism not meeting criteria for Cushing syndrome, and our study raises more questions for future work to answer. Larger studies of populations with varied, but well characterized, cardiometabolic risk factors – at lower risk than the populations in CATALYST and MOMENTUM, such as ours -- are necessary to better estimate prevalence across the cardiometabolic spectrum, as well as to identify clinical predictors to optimally identify patients warranting testing for hypercortisolism.

Though the physiology of cortisol has been studied for 80 years, the causes and long-term implications of mild perturbations remain poorly understood. The direction of causality between cardiometabolic risk factors and hypercortisolism is unclear: is cortisol “the chicken or the egg?” Cardiometabolic risk factors are physiologic stressors that may provoke long-term HPA axis activation, which may in turn over time provoke adrenocortical autonomy. The findings of reversible ACTH hyperstimulability in depression may serve as a model, in which the analogous treatment of the underlying stressor reduces the HPA axis stress response.

### Conclusions

Across cohorts with variable cardiometabolic risk, the rate of ACTH-independent hypercortisolism appears to increase in parallel to risk. Those with ACTH-independent hypercortisolism have similar 8 AM cortisol and ACTH and 24-hour cortisol production compared to those without ACTH-independent hypercortisolism. However, those with ACTH-independent hypercortisolism have higher cortisol after ACTH stimulation. Together, these findings suggest several important avenues for future research into the intertwined pathophysiology of ACTH-independent hypercortisolism, ACTH-induced hypercortisolism, and cardiometabolic risk.

## Data Availability

All data produced in the present study are available upon reasonable request to the authors

